# Use of patient centred outcomes measures alongside the personal wheelchair budget process in NHS England: a mixed methods approach to exploring the staff and service user experience of using the WATCh and WATCh-Ad Tools

**DOI:** 10.1101/2022.08.14.22278761

**Authors:** Lorna Tuersley, Naa Amua Quaye, Rhiannon Tudor Edwards, Nathan Bray

## Abstract

**Background and Objective:** Personal wheelchair budgets (PWBs) are offered to everyone in England eligible for an NHS wheelchair, to support their choice of equipment. The WATCh (Wheelchair outcomes Assessment Tool for Children) and related WATCh-Ad tool for adults are patient centred outcomes measures (PCOMs) developed to help individual users express their main outcome needs when obtaining a wheelchair and rate their satisfaction with subsequent outcomes after receiving their equipment. We explored their use in a real-world setting, aiming to produce guidance for use alongside the PWB process.

**Methods:** Three wheelchair service provider organisations across four sites participated. Staff and users completed surveys about their experience of the WATCh and/or WATCh-Ad tools used in the assessments. Towards the end of the study, selected patients were interviewed after receipt of their equipment, and staff were interviewed after experiencing a number of assessments. Thematic analysis was undertaken using the tool, survey and interview data.

**Results:** Information on 75 assessments by 15 staff was obtained. Over three-quarters of users or their carers rated the use of the tools in the assessment process as ‘helpful’ or ‘very helpful’. Staff reported that use of the WATCh tools had been considered ‘useful’ in developing individual care plans in around 1 in 3 cases and affected the prescription in 1 in 4 cases. Concerns were expressed around the length of time taken to administer the tools in clinic, although some staff noted this reduced with more hands-on experience, and by providing the tools to users in advance of the appointment.

**Conclusions:** The WATCh and WATCh-Ad PCOM tools are suitable for routine use by wheelchair service providers to assist the assessment process. It is recommended that tool materials are provided in advance to users/carers, and that staff are allowed time to develop their ways of working with them.

## Introduction

There are over 14.6 million people in the UK living with an impairment, disability or limiting chronic illness, and almost half of these have a mobility impairment [1]. More than 600,000 people were registered with wheelchair services in NHS England in March 2022, including almost 62,000 children. The average Clinical Commissioning Group (CCG) receives over 100 new and re-referrals for wheelchairs every quarter [2].

Research has highlighted that ineffective provision of wheelchairs increases costs to users in terms of potential harm to health of delayed or ill-fitting equipment, and to the NHS in terms of wasted resources and to society as a whole if users are not fully integrated into society [3].

The NHS has responded to demands to increase the efficiency of service provision and address the needs of the individual users to provide wheelchairs that meet individual needs in a number of ways. These include collection of quarterly data on service provision by CCGs [4]; and the introduction of personal wheelchair budgets (PWB) to help provide everyone with access to a wheelchair that meets their individual health and wellbeing needs and goals [5]. The legal right to a personal health budget of people who access wheelchair services, whose posture and mobility needs impact their wider health and social care needs came into force in December 2019 [6].

The WATCh (Wheelchair outcomes Assessment Tool for <u>Chi</u>ldren) tool was developed as part of an NHS England funded programme of research to develop Patient Centred Outcomes Measures (PCOMs) for use with children and young people (7). The WATCh tool lists 16 pre-determined outcome areas, from which users can select the five outcomes of most importance to themselves at their assessment and rank them in order of importance (Part A). There is also the option to describe an ‘Other’ outcome should they feel there is something important that is not covered in the predetermined list. Users then state, in their own words, what they wish to achieve for each outcome. In Part B, they rate their current satisfaction with these outcomes, using a 5-point scale of ‘smiley faces’, ranging from 1 – ‘Very Unsatisfied’ to 5 – ‘Very satisfied’. Some months after receipt of the equipment, a follow-up tool, Part C, is sent to the user to complete, requesting a re-rating of their satisfaction against their chosen outcomes. Ongoing work on the quality of life of users of mobility equipment by the authors has shown that the general outcomes desired by adults are closely related [8], hence the wording of this tool was adapted for use with adults as WATCH-Ad. The tools can be used as paper based (See Supporting information 1) or can be completed electronically (see https://cheme.bangor.ac.uk/watch.php).

The uptake of any new treatment or process relies on successful implementation. NHS England have sought to embed PCOMs into the PWB pathway, and funded the research described to help develop guidelines for roll-out nationwide. Our aim was to assess outcome achievement at the level of the user and identify aspects important to them and the service provider, in order to maximise wider implementation. A further aim was to investigate the practicalities of use and the resource implications of introducing a PCOM into the PWB process.

## Methods

A mixed methods approach was used, comprising staff and service user survey questionnaires, telephone interviews and analysis of the completed WATCh and WATCh-Ad tools. The researchers worked with the NHS England personal health budgets team and the National Wheelchair Advisory group (NWAG) which includes wheelchair service providers and users, to develop the scope of the work and inform the questionnaires and interview schedules. A study management team involving lead contacts from the participating organisations met throughout the study to oversee progress. The study was approved by Health and Care Research Wales (20/WA/0007).

In November 2019, NHS England sent expression of interest invitations (EoI) to wheelchair services supporting CCGs. Participating sites were selected based on obtaining a mix of experience of use of the WATCh and or WATCh-Ad tools and the implementation of PWBs, geographic location and whether they were NHS-staffed or managed through independent contractors. The four sites from three provider organisations had differing levels of experience: one with prior experience of both the PWB process and of the tools, provided through an independent contractor in the North East of England (Site A), two smaller sites in the Midlands run by a second independent contractor organisation, with prior experience of the PWB process but not of the tools (Sites B and C), and one provided directly though the NHS in the South East with no experience of either (Site D). Table 1 shows the information provided in response to the EoI by the selected organisations.

**Table 1:** Participating Organisations and Service sites.

| SITE | A | B | C | D | B/C* | TOTAL<br>A, B/C,<br>D |
| --- | --- | --- | --- | --- | --- | --- |
| LOCATION of Service | North of<br>England | Midlands | Midlands | South<br>East | Midlands |  |

| NHS/Independent contractor |  | Independent | *Independent* | *Independent* | NHS | Independent |  |
| --- | --- | --- | --- | --- | --- | --- | --- |
| Start use of PWB |  | Feb-19 | *Apr-19* | *May-19* | Dec-19 | Yes |  |
| Start use of WATCH tools |  | Feb-19 | - | - | - | - |  |
| Adults/<br>quarter | New | 425 | *206* | *119* | 530 | 325 | 1280 |
|  | Re-referrals | 545 | *283* | *132* | 356 | 415 | 1316 |
| Children/<br>quarter | New | 33 | *21* | *18* | 90 | 39 | 162 |
|  | Re-referrals | 110 | *52* | *64* | 79 | 116 | 305 |
| Total Adults |  | 970 | *489* | *251* | 886 | 740 | 2596 |
| Total Children |  | 143 | *73* | *82* | 169 | 155 | 467 |
| Total New |  | 458 | *227* | *137* | 620 | 364 | 1442 |
| Total re-referral |  | 655 | *335* | *196* | 435 | 531 | 1621 |
| TOTAL |  | **1113** | ***562*** | ***333*** | **1055** | **895** | **3063** |

\*Sites B/C combined as managed by same organisation and similar overall users to A and D

The study sought to obtain information from staff and service users on their views of the assessment process using the WATCh tools and the PWB process. In addition, telephone interviews were to be carried out among a range of service users following receipt of their new wheelchair, and with service staff who had experience of working with the tools.

Participation packs for staff and potential participants were provided in advance, including information about the study, consent or proxy forms, the WATCh or WATCh-Ad tools and the staff or participant survey forms (See Supporting information 2). Information sheets were provided according to age group; under 6 years, 6-10 years, 11-15 years and 16 and over (adults). After the assessment, staff and the users or their carers completed short survey questionnaires about their experience of the use of the tools in practice.

Staff were asked to record the time taken by the service user or carer to complete the tool and who completed it. They were also to note if any difficulties were experienced and whether the staff member was able to resolve them. They were invited to comment on usefulness of the tool, any impact it had on the prescription and the PWB option selected.

The user survey questionnaires asked service users or carers for more personal details about the user, their need for a wheelchair and the time since their previous assessment (if any). They were asked about the practicalities of completing the tool including time to complete, how easy it was to understand, any questions they had and whether they felt that any outcome areas were missing. Consent was sought for information about their assessment and any follow up data being made available to the research team in an anonymised format. Contact details were only required if they consented to a follow-up interview. Where a user was considered by the assessor site to be unable to complete responses due to age, capacity or health issues, their parent/guardian or carer was invited to submit responses and complete consultee forms as appropriate. Staff and user surveys were handed in to the clinic co-ordinator in sealed envelopes for return to the researcher, to protect the anonymity of the responses.

The WATCh and WATCh-Ad tool data was shared with the researchers to allow them to quantify the outcomes selected and their scores, and any changes between the assessment and follow-up outcomes. The qualitative data on reasons for choice of outcomes was also reviewed. Given the short timescale for this study, it was anticipated that not all users involved in the study would obtain and gain experience of their equipment to be able to complete the follow-up WATCh tool. Thus, staff in sites with prior use of the tool were asked about their experience of follow-up during their interviews. In addition, such sites were asked to provide anonymised, non-identifiable data from prior assessment and follow-up scores for descriptive statistical analysis.

A number of staff and service users were invited to be interviewed over the telephone, exploring their responses to the survey and the use of the tools in the PWB process. These included a manager and therapists from each organisation; and purposive sampling of service users representing those having their first assessment, those who had previously been given a wheelchair, adults and children and whether the tool was completed by the user or their parent/carer. Consent was obtained in advance of data collection.

### Impact of the pandemic upon the methods

On the 19th of March 2020, the Director of Community Health for NHS England and NHS Improvement wrote to providers of community services, directing them as to how capacity could be released to support the COVID-19 preparedness and response [9]. This included a significant reduction in wheelchair clinic provision. Due to staff redeployment, Site D was unable to start recruitment before this date. In addition, all non-COVID-19 related research was paused during this time period. Permission was given by funders, and the University’s School ethics approval and local Research Ethics committee (L-REC) to extend the study until 31^st^ March 2021. At the end of May 2020, the National Institute for Health Research (NIHR) issued guidance on the Restart Framework [10] and in July 2020, activities could be re-started at the sites contracted to independent organisations, who had previously recruited participants. Site A continued to triage patients prior to attending clinic, but it was agreed that Sites B and C would cease recruitment as sufficient numbers had been identified elsewhere. Staff at site D were able to start planning for recruitment, following review with their organisations’ research and development departments. Updated information for adults relating to General Data Protection Regulation requirements (GDPR) were approved by the authors’ institution and ethics committee in December 2020 to allow interviewing to start.

In January 2021 it was agreed that data collection would be cut off on 16^th^ February, to allow for review of data and follow up of any queries within the extension period. At site D, although 24 assessments were planned during this period, use of the tools was only possible in 19. Five were deemed unsuitable by the clinician as they were home visits for vulnerable, shielding patients, where contact time needed to be kept to a minimum.

Assuming a consent to interview rate of 10%, the protocol had stated that each organisation should aim for a total of 100 assessments to be carried out. Although the number of participants was much lower than this due to the pandemic, the consent rate to interviews was over 70% and represented the majority of the participant types required.

Despite the very high consent rate to interviews given in the patient surveys, of the first six consenting users contacted, only two interviews subsequently took place. Two did not respond to phone calls, one declined to take part, and one interview was cancelled due to family circumstances due to the pandemic. The two interviews that did take place gained limited additional information compared with the users’ surveys. Given this low uptake and that the delay since the original assessment in most cases would require recall of quite distant events, it was agreed that no further attempts would be made to contact users. Consents from staff were high and six interviews with staff took place as planned, which were able to provide additional information to their surveys based on information about use in practice and their reflections on this. Research activity on the study stopped at the end of March 2021 in line with the deadline to the extension.

### Analysis

Data from the service users’ surveys and their associated staff survey was entered into Microsoft Excel for each service user. The datasets were explored and analysed for descriptive statistics including by subgroups of site, user versus staff reports, adults versus children and first-time assessments versus re-referred patients. Percentages are based on 75 participants, the total of assessments completed by staff. In five cases the patients themselves did not want their survey data included so have been included as ‘not stated’ (n/s) responses.

Qualitative data obtained through the surveys and the interviews was transcribed and analysed for emerging themes assisted by use of a Microsoft Excel spreadsheet.

## Results

### Numbers recruited

Information on a total of 90 potential uses of the tools in assessments was returned by the sites: 27 from Site A, 39 from sites B and C combined, and 24 from Site D. Information on ten planned assessments at Site C could not be included as five users or their carers did not want to take part; two felt the survey and tool were too long, two appointments were cancelled and one failed to provide any written consent to the study. Five potential uses in home visits at site D did not take place due to the user being considered vulnerable and was shielding.

The data available from 75 completed assessments is shown in Table 2.

**Table 2:** Data returned by Organisation.

| **SITE** | **Staff survey**<br>Total *n* (children) | **User survey**<br>Total *n* (children) | **Tool data**<br>Total *n* (children) |
| --- | --- | --- | --- |
| A | 27 (2) | 27 (2) | 26 (2) |
| B/C | 29 (10) | 26 (10) | 26 (9) |
| D | 19 (10) | 17 (9) | 15 (8) |
| **TOTAL** | **75 (22)** | **70 (21)** | **67 (19)** |

A total of 75 staff surveys were returned by 15 members of staff. User surveys were completed by 70 users or their parents/carers and 67 completed WATCh assessments were obtained (19 WATCh and 48 WATCh-Ad)

### Staff

Surveys were returned by 15 different staff including ten occupational therapists (OT), two OT/Clinical leads, one physiotherapist (PT), and one research engineer (RE) (Table 3). One did not state their occupation. Interviews were carried out with 6 staff between December 2020 and February 2021.

**Table 3:** Number of surveys and interviews completed by wheelchair service provider staff at each organisation

|  | <u>SITE A</u> | <u>SITE B/C</u> | <u>SITE D</u> | <u>TOTAL</u> |
| --- | --- | --- | --- | --- |
| OT *n* (*n* surveys) | 5 (22) | 4 (22) | 1 (1) | 10 (45) |
| OT Interview |  | 2 |  | 2 |
| PT *n* (*n* surveys) |  |  | 1 (18) | 1 (18) |
| PT interview |  |  | 1 | 1 |
| Manager (*n* surveys) | 1(5) | 1(1) |  | 2 (6) |
| Manager Interview | 1 | 1 |  | 2 |
| Other *n* (surveys) |  | 2 (6) |  | 2 (6) |
| Other Interview | 1 |  |  |  |
| TOTAL Staff (*n* surveys) | 6 (27) | 7 (29) | 2 (19) | 15 (75) |
| TOTAL interviews | 2 | 3 | 1 | 6 |

### Service Users

Details of participants are provided at Supporting information 3. Information was returned relating to 75 assessments in total, 53 adults and 21 children aged under 16, with ages ranging from 2 years to 90. Table 4 shows the break down by age and gender for each site.

**Table 4:** Age and sex of service users by wheelchair service site.

|  |  |  | *N* | Mean age (years) | Range (years) |
| --- | --- | --- | --- | --- | --- |
| SITE A | Adults | Male | 11 | 45.8 | 28-73 |
|  |  | Female | 14 | 51.1 | 19-86 |
|  | Child | Male | 1 | 7.0 | 7.0 |
|  |  | Female | 1 | 6.0 | 6.0 |
|  |  | TOTAL | 27 | 45.6 | 6-86 |
| SITE B/C | Adults | Male | 10 | 59.3 | 27-77 |
|  |  | Female | 6 | 48.8 | 22-90 |
|  |  | n/s* | 3 | 76.3 | 68-81 |
|  | Child | Male | 5 | 7.6 | 2-13 |
|  |  | Female | 4 | 8.0 | 2-15 |
|  |  | n/s* | 1 | 5.0 | 5.0 |
|  |  | TOTAL | 29 | 40.9 | 2-90 |
| SITE D | Adults | Male | 3 | 56.7 | 44-76 |
|  |  | Female | 4 | 49.8 | 18-71 |
|  |  | n/s* | 2 | 42.5 | 28-57 |
|  | Child | Male | 4 | 9.5 | 5-15 |
|  |  | Female | 4 | 7.5 | 5-13 |
|  |  | n/s* | 2 | 10.5 | 9-12 |
|  |  | TOTAL | 19 | 28.1 | 5-76 |

Of those reporting, 8 adults and 12 children were first time attendees to wheelchair services, 11 adults and 9 children had been previously seen by a wheelchair service within the past 12 months, 12 adults and 4 children had been seen by a wheelchair service between 1 and 5 years ago, and 9 adults had been seen longer than 5 years ago.

Service users’ reasons for requiring wheelchair services were varied and many only provided general statements relating to difficulty getting around. Table 5 shows the range of underlying reasons split by adults and children and by whether this was their first assessment or not.:

**Table 5:** Underlying need for wheelchair reported by new or returning user.

|  | First-time wheelchair service user |  | Returning wheelchair service user |  | First or returning n/s |  |
| --- | --- | --- | --- | --- | --- | --- |
|  | Reason | *n* | Reason | *n* | Reason | *n* |
| ADULTS | General | 4 | General | 10 | n/s | 6 |
|  | n/s | 2 | Multiple Sclerosis | 3 | Cerebral Palsy | 1 |
|  | Amputee | 1 | Cerebral Palsy | 3 | Frederick ataxia | 1 |
|  | Arthritis | 1 | Amputation | 2 | Multiple Sclerosis | 1 |
|  | Falls | 1 | Pain | 2 |  |  |
|  | Fibromyalgia | 1 | Arthritis | 1 |  |  |
|  |  |  | Arthrogryposis/Multiple |  |  |  |
|  | Parkinson's | 1 | sclerosis | 1 |  |  |
|  | Spinal condition | 1 | Broken hips | 1 |  |  |
|  |  |  | Ehlers Danlos Syndrome | 1 |  |  |
|  |  |  | Huntington's | 1 |  |  |
|  |  |  | Myotonic Dystrophy | 1 |  |  |
|  |  |  | n/s | 1 |  |  |
|  |  |  | Nerve damage | 1 |  |  |
|  |  |  | Paraplegia | 1 |  |  |
|  |  |  | Spinal arthritis | 1 |  |  |
|  |  |  | Stroke | 1 |  |  |
|  |  |  | Stroke/Arthritis | 1 |  |  |
|  | **TOTAL** | **12** | **TOTAL** | **32** | **TOTAL** | **9** |
| CHILD | Autism/ global |  |  |  | n/s | 1 |
|  | development | 1 | General | 8 |  |  |
|  | Cerebral Palsy | 1 | Cerebral Palsy | 3 |  |  |
|  |  |  | Dandy-Walker |  |  |  |
|  | Complex | 1 | Syndrome | 1 |  |  |
|  | General | 1 | Hypermobility | 1 |  |  |
|  | Hypermobility | 1 |  |  |  |  |
|  | Pain | 1 |  |  |  |  |
|  | Seizures | 1 |  |  |  |  |
|  | n/s | 1 |  |  |  |  |
|  | **TOTAL** | **8** | **TOTAL** | **13** | **TOTAL** | **1** |

Despite the smaller number of participants than originally planned, there was a high rate of consent to interview, given by 54 users or their carers (70%). These represented most of the respondent types required across all sites, by age, site, whether new or returning user or the tool was completed by user or carer, with the exception of children under 16 completing the tools themselves (Table 6).

**Table 6:**
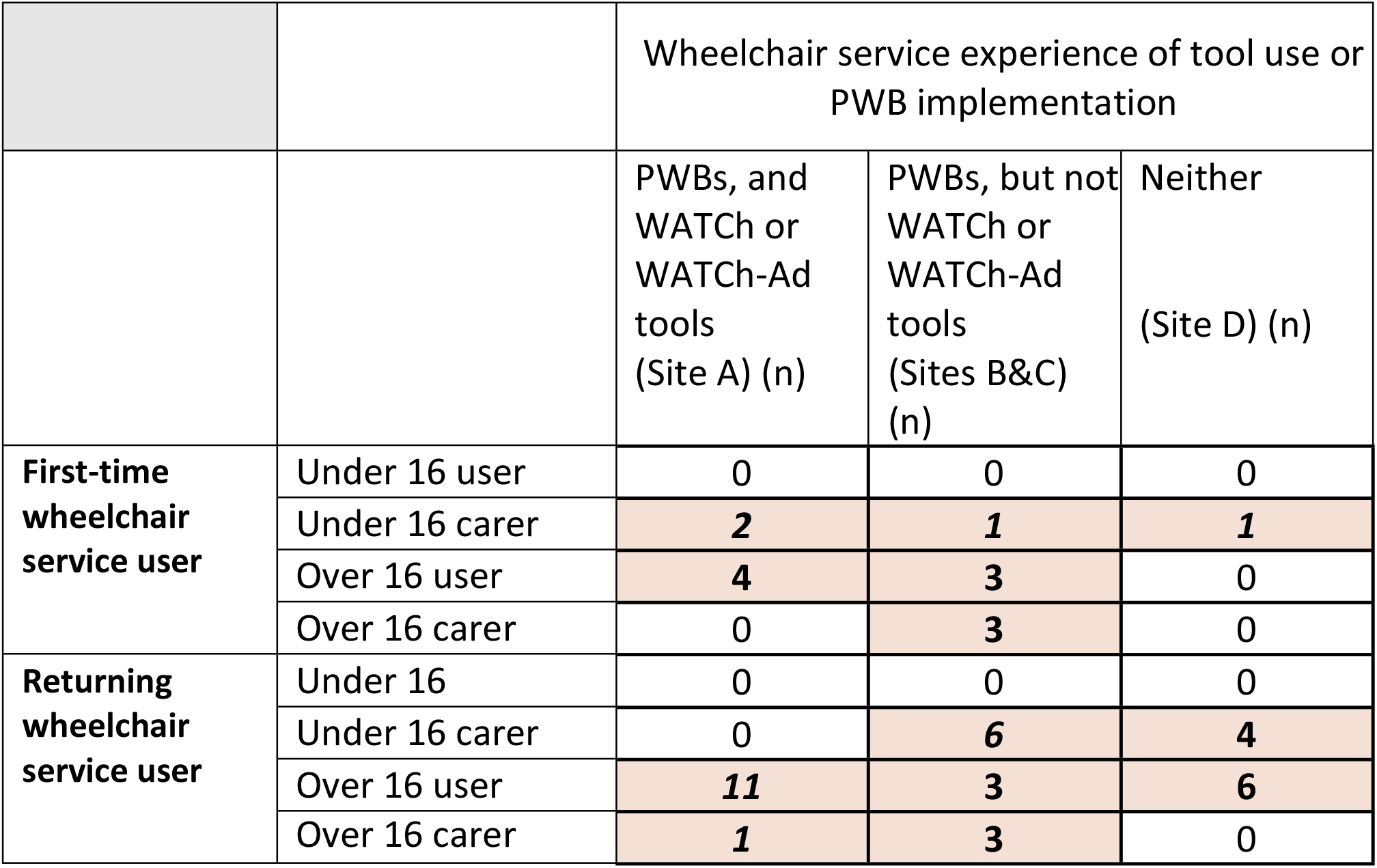
Number of service users/carers consenting to take part in an interview.

### Survey findings

Staff completed a survey a total of 75 assessments carried out using the tools, asking about use of the tools in practice, including questions about any additional time taken compared to an assessment without using them. They were specifically asked about any problems with service users’ ability to complete the tool and their own ability to deal with any issues arising. These were compared with the user survey responses to similar questions.

The mean time taken to complete the form was reported to be 12.2 minutes by staff and 13.1 minutes by users (Fig 1)

**Fig 1:**
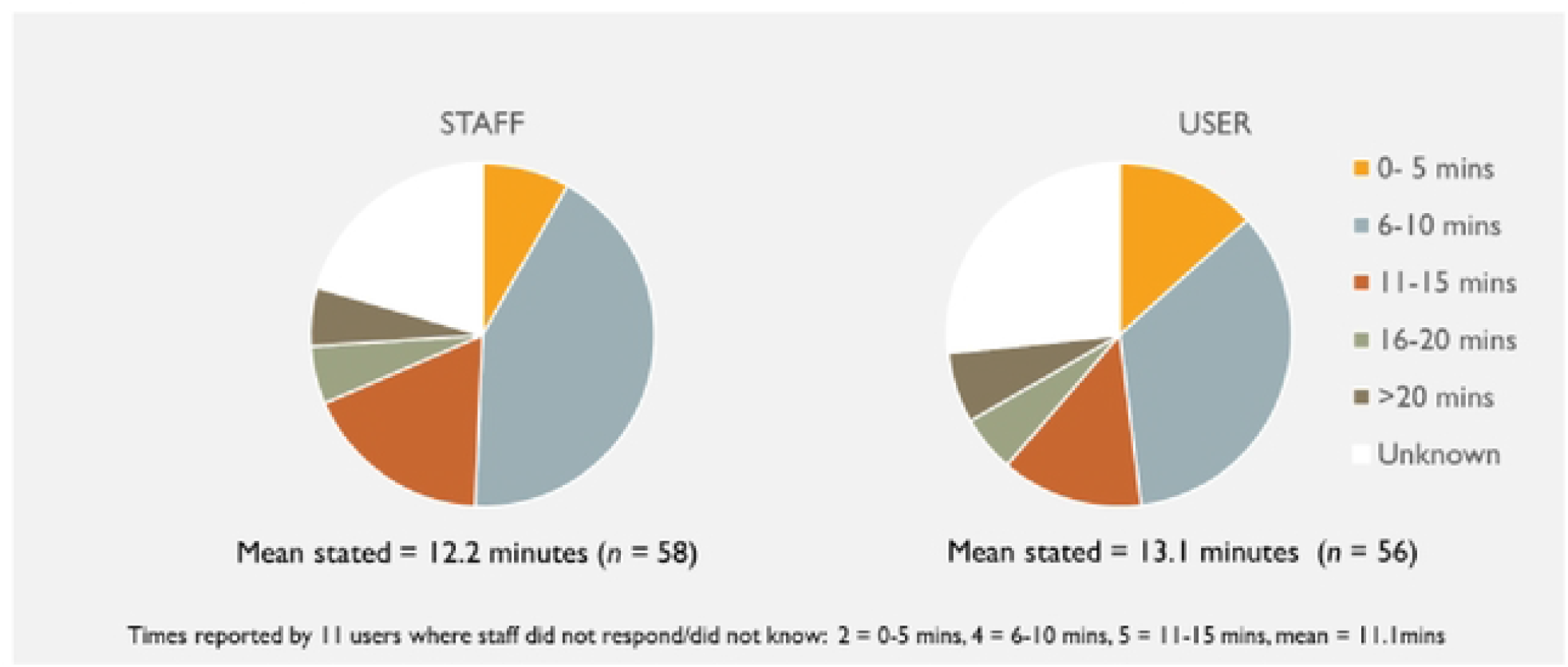
Time to complete WATCh or WATCh-Ad tool – Staff versus user surveys.

In terms of the person completing the tool forms, in over two-thirds of assessments this was stated to be the user or carer without help from staff. (Figure 2).

**Fig 2:**
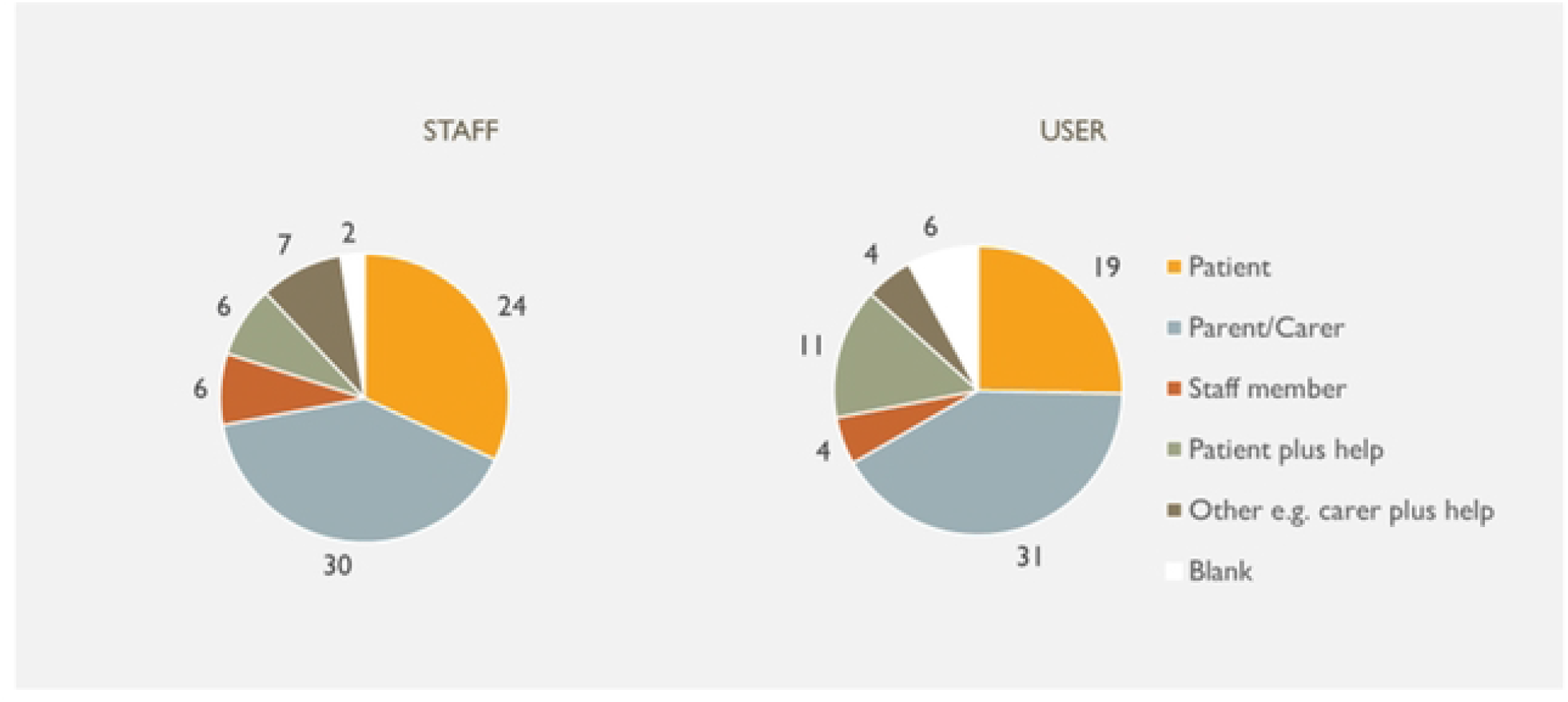
Person completing the WATCh or WATCh-Ad tool.

Staff were asked about any problems with completing the tools and whether they were able to deal with them, as Yes/No options (Fig 3). In 35 (47%) cases, no problems were reported. Comments were made by 19 giving ‘No’ or n/s responses. The problems described related to inability to complete without explanation (*n* = 5), administration of the form, for example omitting to bring to clinic or client felt rushed (*n* = 5), difficulty in choosing outcomes or setting goals (*n* = 4) and communication issues such as dyslexia, writing difficulties and need for family member to translate (*n* = 3). In one case the staff member felt it was irrelevant as the service user only needed adjustment to the headrest and in another, the parent of a child with a terminal condition found the tool to be insensitive.

**Fig 3:**
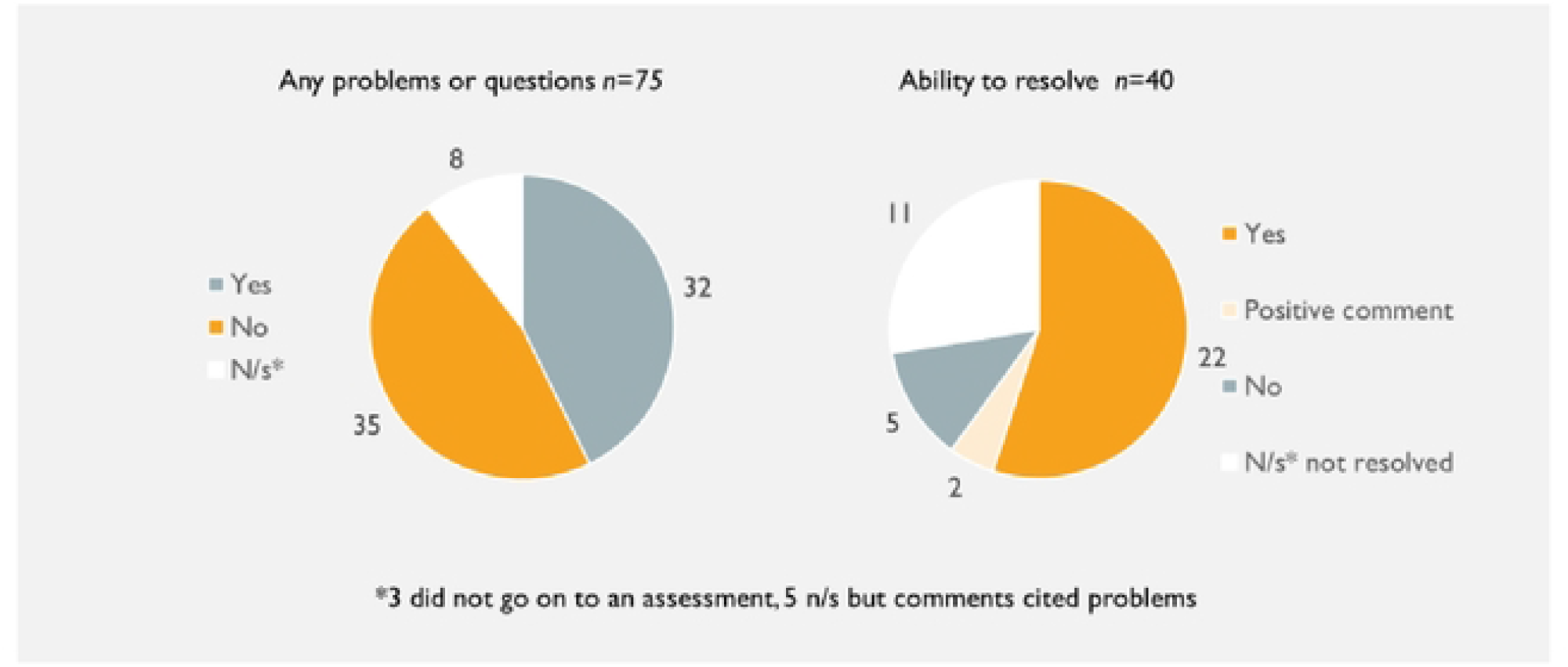
Staff noting user problems or questions with using the tools and ability to resolve.

In contrast, the majority of users or carers (n=61; 81%) felt that it was easy to understand and 75% (n=56) reported no problems with using the tools. Comments from seven who felt it was not easy to understand, included that it was too long or not relevant for what they were being assessed for, and one because they completed it over the phone. Five responded ‘yes’ to the question about whether anything was missing; where made, comments related to the general approach for of the tool or to the user’s complex clinical condition, rather than their expectations from a wheelchair. See Figure 4.

**Fig 4:**
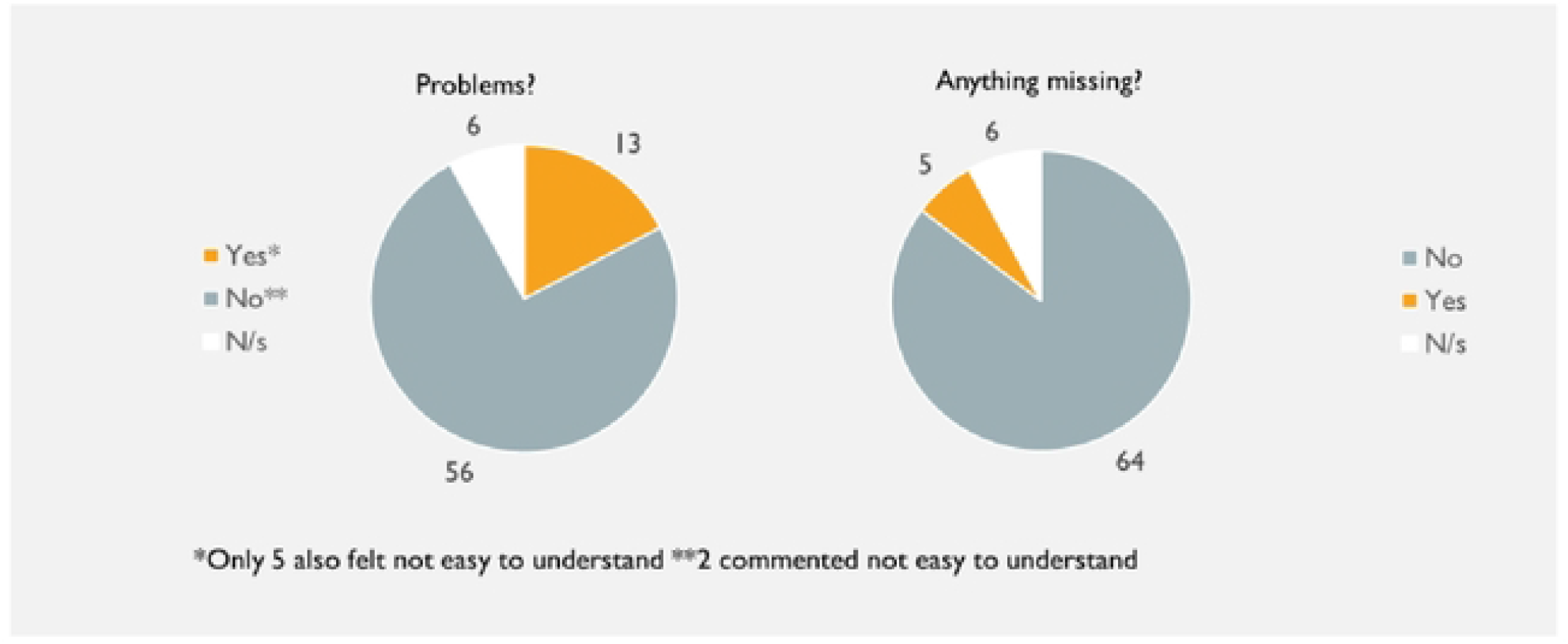
User or carer view of problems with the WATCh or WATCh-Ad tool.

Staff were also asked whether they assessed use of the tools as useful or not. They were stated to be useful in 35% (*n*=26) of assessments but not useful in 41% (*n*=31). In contrast, when users were asked to rate helpfulness on a scale from ‘very helpful’ to ‘not helpful at all’, 45% (*n* =34) rated the tool as very helpful and a further 32% rated it as quite helpful. (Fig 5).

**Fig 5:**
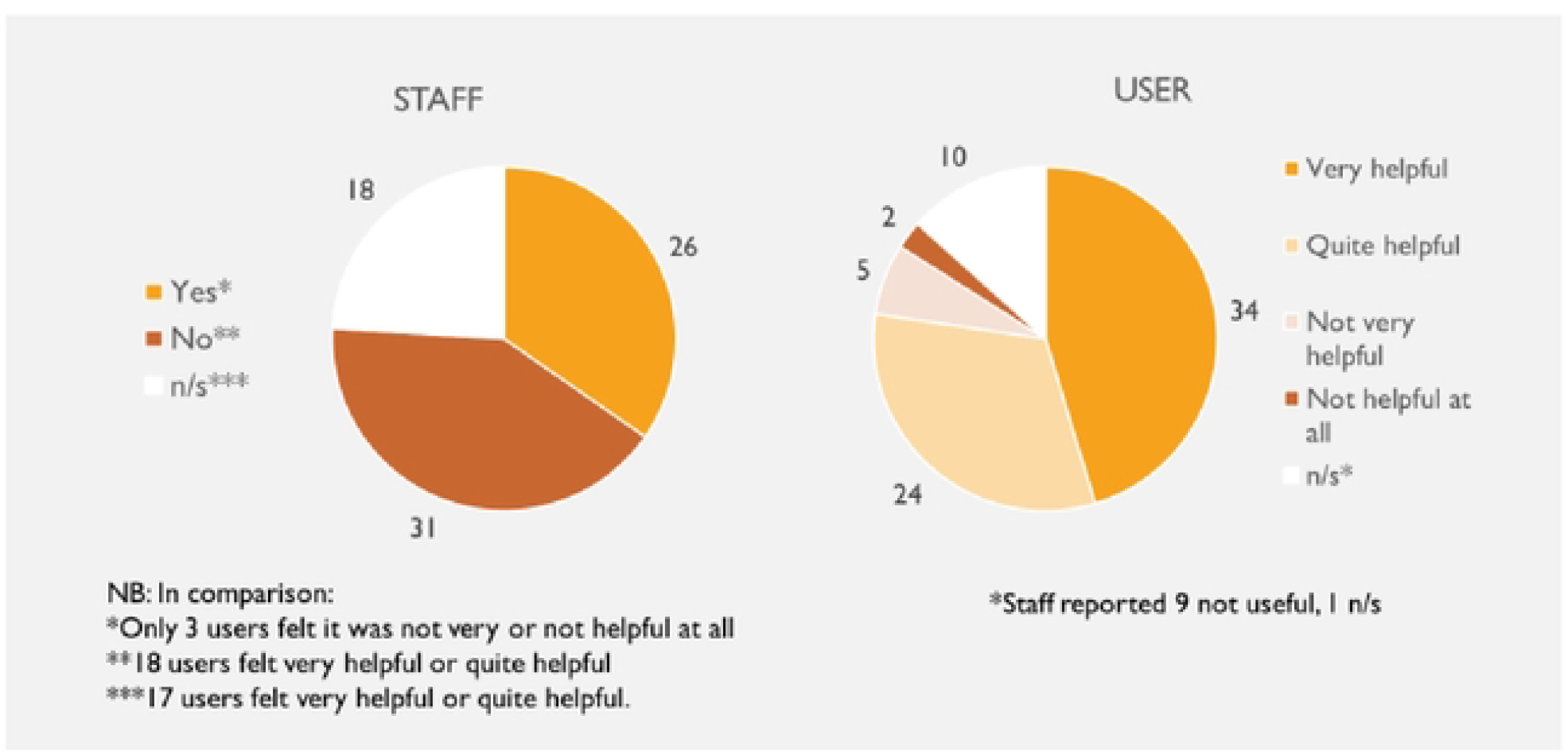
Comparison of staff and user rating of usefulness and helpfulness.

Positive responses from staff included that it was good for highlighting areas of importance or difficulties being faced by the service user and/or the caregiver; for directing the reasons for the assessment and allowing a more in-depth review of an individual’s outcomes. Reasons for negative responses included considering that it had not influenced their practice as their clinical requirements would have been provided anyway; that the goals had already been discussed, or that the client was well-known to staff. In some cases a specific practical aspect such as the child having outgrown the chair or that a footplate needed adjustment was not picked up despite ‘comfort’ being an option for selection on the tool. Some expressed concerns that the goal setting might be biased if the client had difficulty in doing this independently, and also the possibility that use could raise expectations.

Staff were asked to report whether using the tool had impacted on their eventual choice of prescription. Fig 6 shows the proportion of staff reporting this for adults and for children.

**Fig 6:**
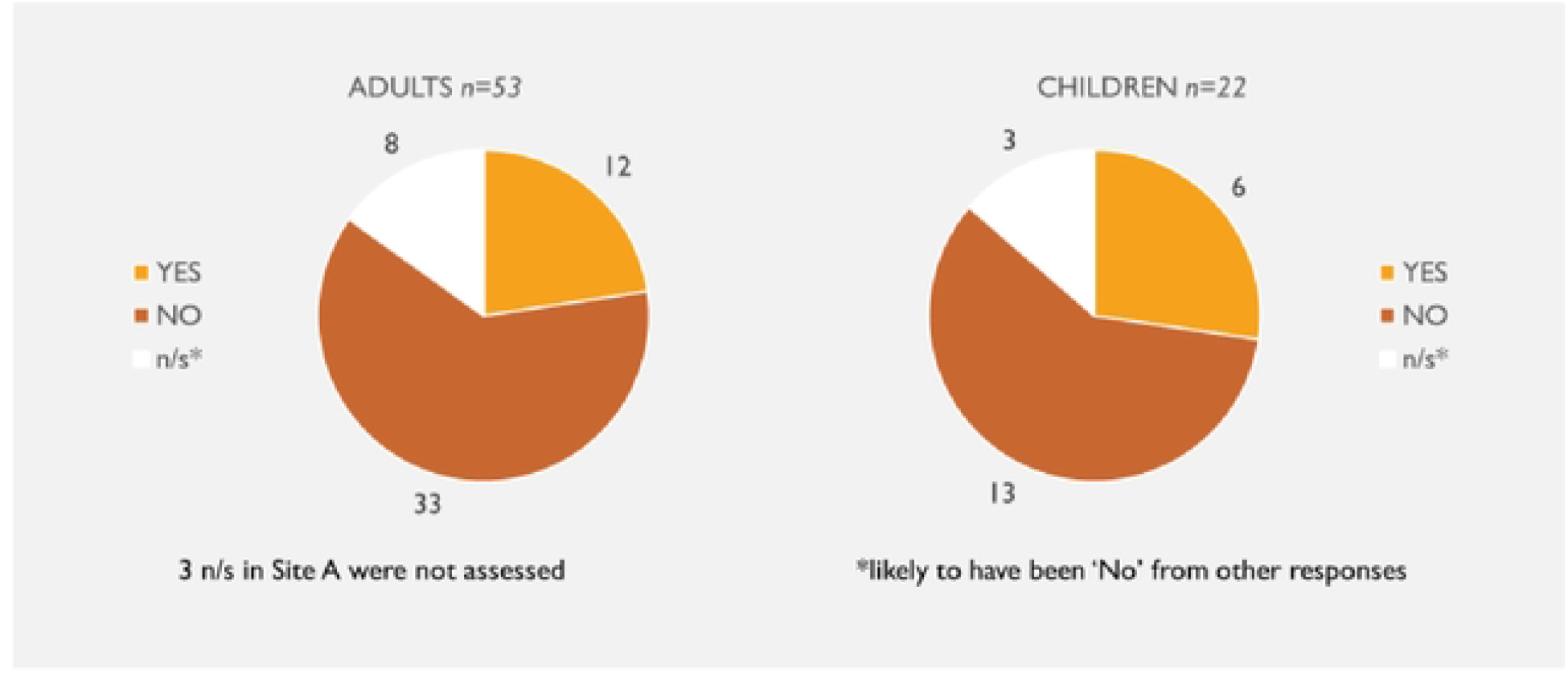
Staff perceptions of the impact of using the WATCh or WATCh-Ad tool on their prescription by cases – Adults versus children under 16.

In around a quarter of the assessments made by staff, it was felt that use of the tools affected the eventual prescription choices: in 23% (12 of 53 assessed) of adults and 27% of the children assessed (6/22). Where an impact was noted, reasons included the choice of power pack, the type of seating, a specific power chair, height adjustable handles for carer needs and the weight of the chair.

> *“At the moment client is using S/P [self-propelled] manual W/Ch [wheelchair] but client will be limited with his independence and will impact his goal to have work and be pain-free and might develop worst pain on wrist, back and shoulder if not given a powered chair” (Site A, adult)*
>
> *“Added height adjustable handles due to mother’s own health issues” (Site D, child)*

The main reasons stated for a lack of impact included that the tools had little impact on clinical decisions or led to the same outcomes as by the usual process or that any changes required were better identified through other means. A limited choice of wheelchair was also mentioned especially in complex cases or due to other factors.

> *“It was not possible to meet the health and wellbeing plan due to the criteria linked to ‘Active wheelchair XXXX’ for people who require a wheelchair for outdoor use only” (Site A, adult)*
>
> *“There are a limited number of buggies available which meet the SU’s [service user’s] clinical needs therefore this had little impact on the clinical decision made*.*” (Site B, child)*

However there was also acknowledgment that the tool supported the prescription decision, even if unchanged

> *“Helped to direct reason for the assessment although same outcome would have been achieved” (Site B, adult)*

Among the users and their carers, positive comments included that the tools made it easier to explain needs to staff, allowed more time to think (if provided in advance) and encouraged everyone involved to think about things not previously considered.

> *“To realise what you actually wanted. Things on there that I didn’t think of” (Site A, adult)*
>
> *“Made it easier to explain as clinician had a rough idea before appointment of my needs without having to explain lots of information. Completing it at home made it easier to fill in with time*.*” (Site A, adult)*

Finally, staff were asked to state if use of the tool had made any difference to the amount of time taken in the assessment to use the tool, both for themselves alone and including any other staff involved. On average, the staff member completing the form estimated it added an extra 11.1 minutes for them to complete the tool, increasing to an extra 16.5 minutes of staff time in total where others were included. (Fig 7).

**Fig 7:**
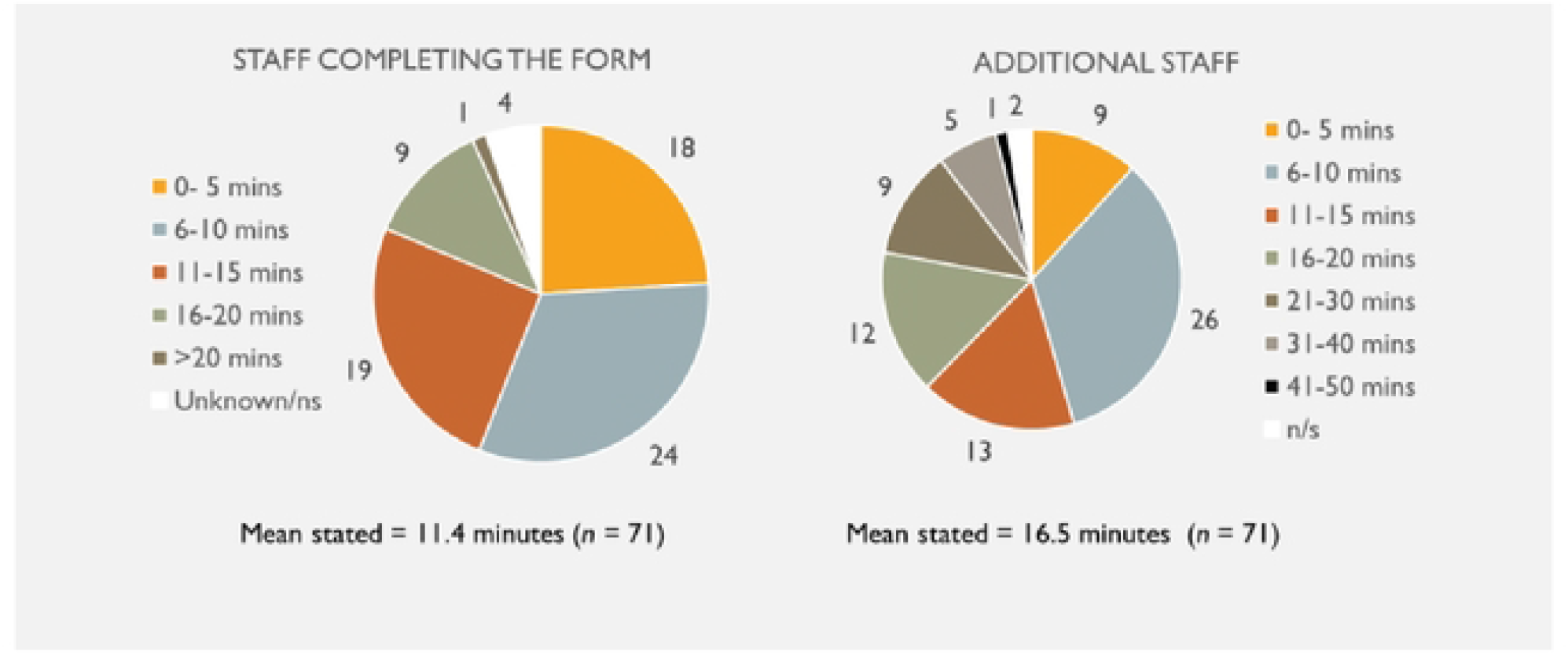
Additional time spent by staff using the tools in the assessment – assessor only and including any additional staff.

When looking at the eventual choice of how the equipment was to be financed, in the majority of cases (68%, *n*=51)) this was stated to be notional NHS provision for both adults and children. For six adults (4 from site A and 1 each from sites B and D), a ‘notional plus contribution’ option was selected and one adult from Site B selected the 3^rd^ party option. (Fig 8). In the latter case, the assessor felt it was the PWB paperwork that had assisted the selection of goals. For those selecting notional plus contribution, in three cases (one each from A B and D) the assessor stated that the prescription had not been affected by use of the Tool any more than their standard practice. In one service (Site A) it was felt that expectations of what could be provided under the notional financing were raised but could not be met. In two cases from Site A, the tool had been useful and had impacted on the prescription.

**Fig 8:**
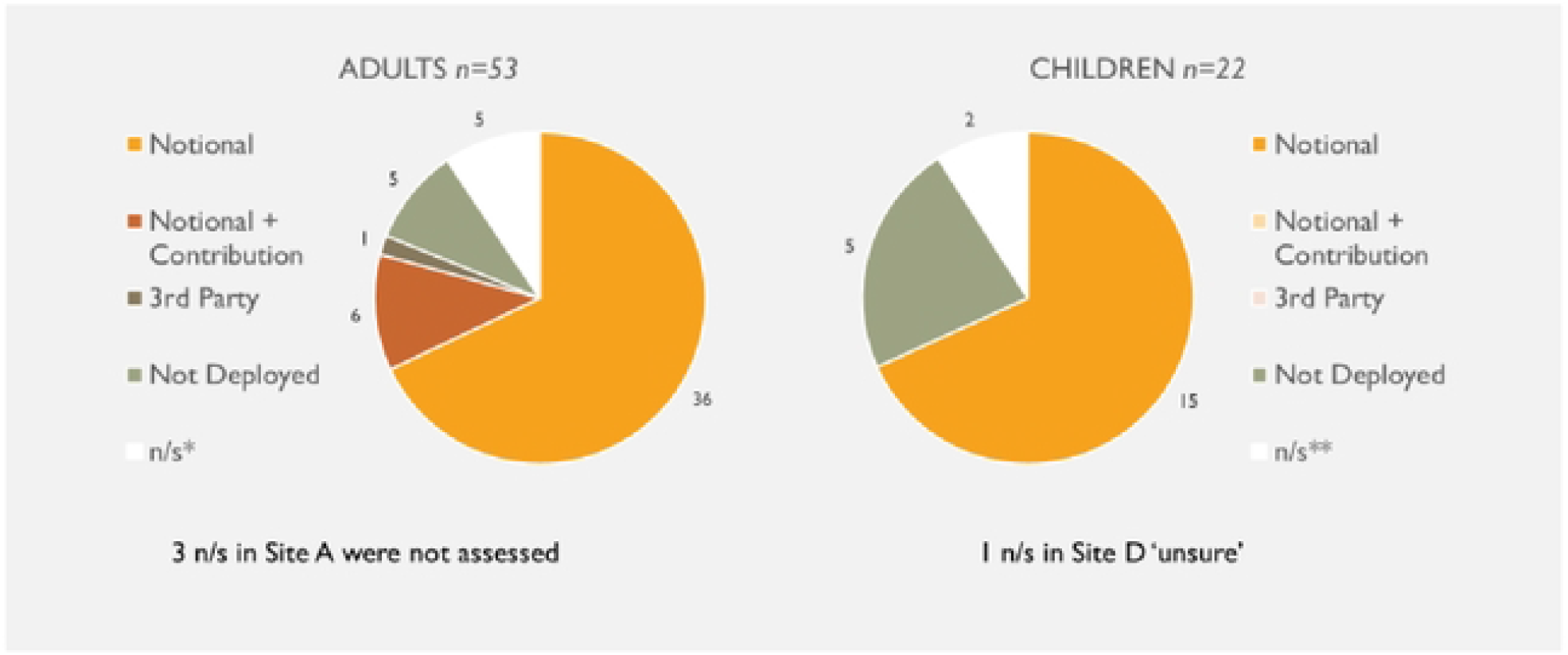
Equipment PWB option selected.

### Tool data

Consent to review their tool data was given by 67 service users or their carers, including 19 children. This provided an opportunity to assess the relative frequency of use of each item and to check that all were relevant.

All items on the WATCh-Ad tool for adults were selected by at least one respondent, including the blank option ‘other’. For the WATCh tool, all items except for ‘Self-care’, ‘Happiness’ and ‘Achievement and goals’ were selected at least once

The most commonly selected items for adults were: ‘Moving around’, ‘Independence’, ‘Pain & discomfort’, ‘Social life’ and ‘Safety’. For children, the most commonly selected were ‘Moving around’, ‘Pain & discomfort’, ‘Social life’, ‘Activities & fun’ and ‘Education’. Figs 9 and 10 show the choices and relative rating by their order on the tool form.

**Fig 9:**
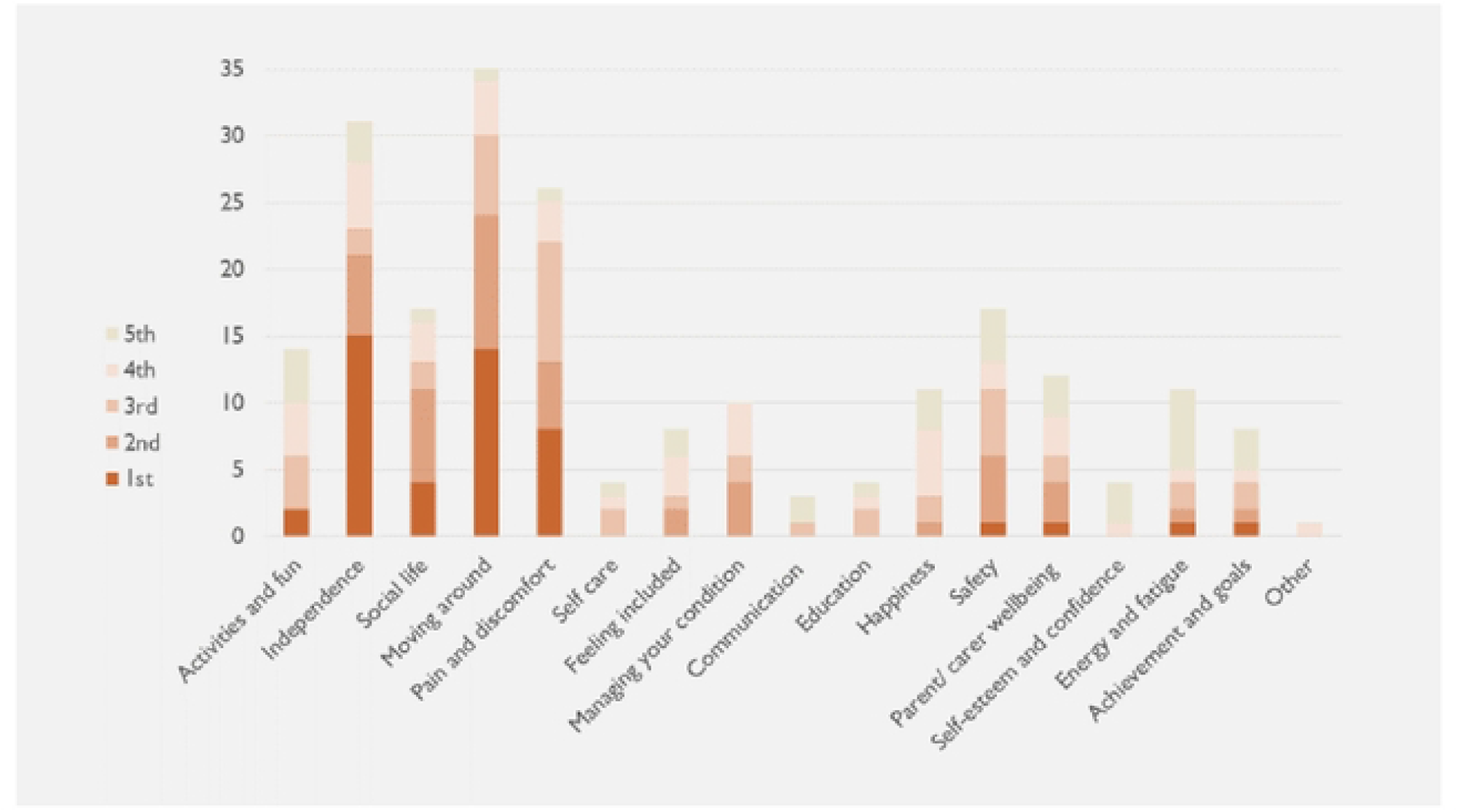
Selection of WATCH-Ad tool outcomes by adults.

**Fig 10:**
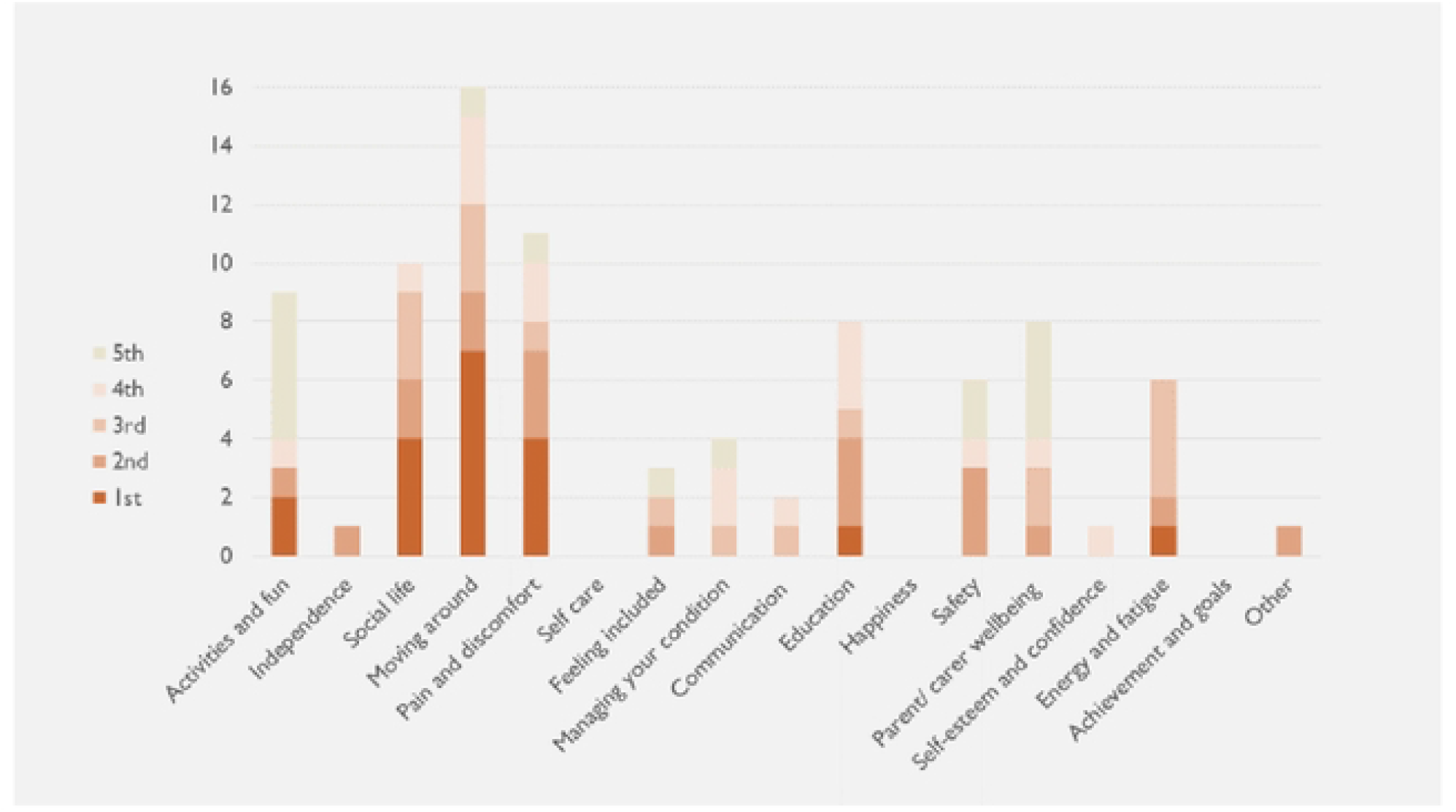
Selection of WATCH-Ad tool outcomes for children.

Boxes 1 and 2 give examples of statements made. Reviewing the users’ description of goals highlighted there was overlap between areas e.g. ‘moving around’ might pick up safety, activities etc.

#### Box 1: Reasons for choice of top outcome item (satisfaction level): Adults

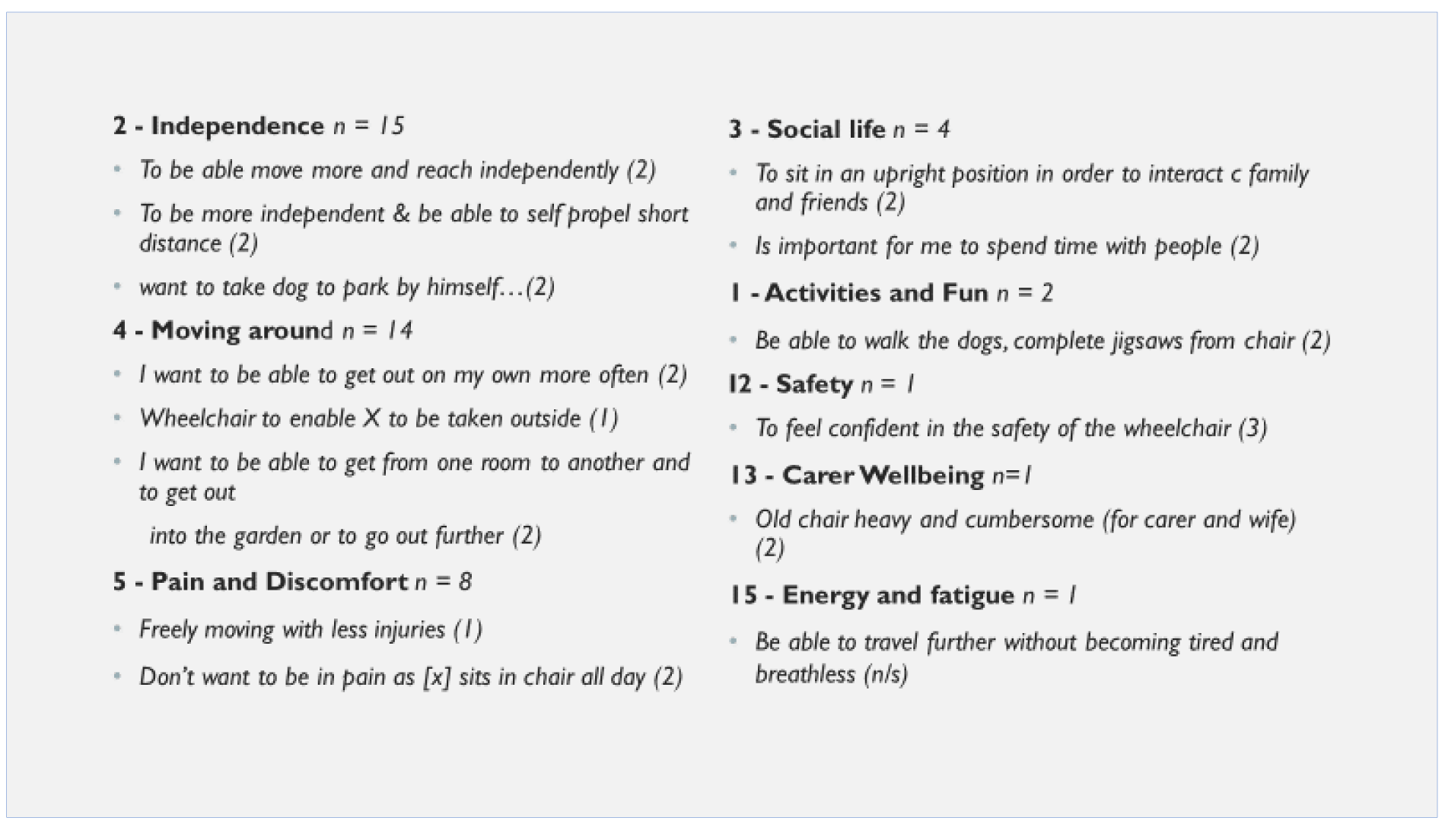

#### Box 2: Reasons for choice of top outcome item (satisfaction level): Children and Young People

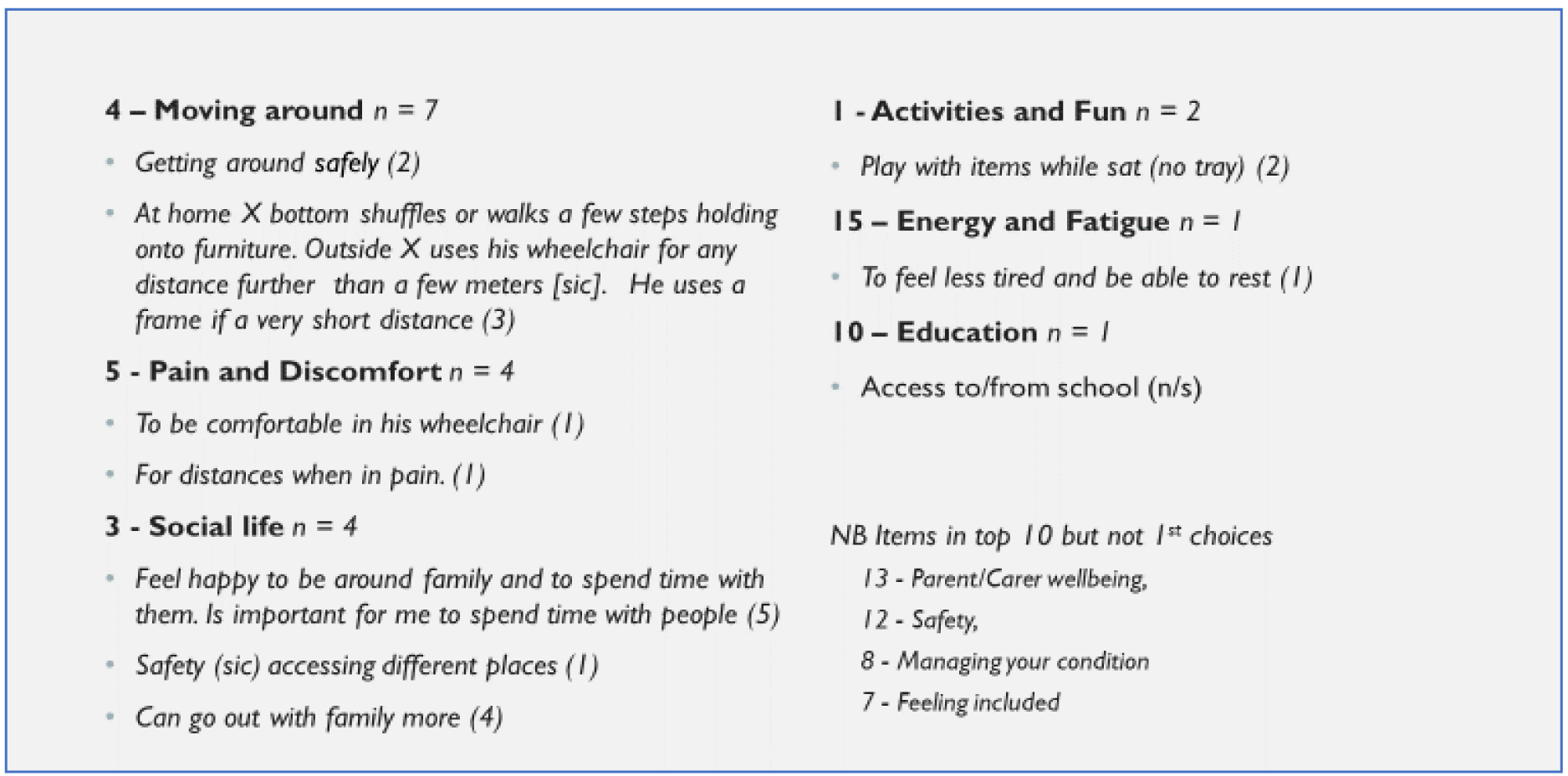

### Follow-up data

The impact on study timings due to the COVID-19 pandemic meant that it was only possible to obtain follow-up data from a small number of respondents at Sites A and B. At site A, 15 of 26 (58%) participants had been provided with equipment, eleven of whom (73%) had provided follow up scores. At site B, eight (53%) of 15 participants available for follow-up provided follow up scores, of which seven had completed formal consent. Assessment and follow-up scores for these eighteen are presented in Fig 11. Where no follow up data was available, this was still being sought or the user had not yet received their equipment. Reasons for the latter included not having decided on a PWB option, lack of availability of equipment or delays in attendance due to COVID-19 isolation.

**Fig 11:**
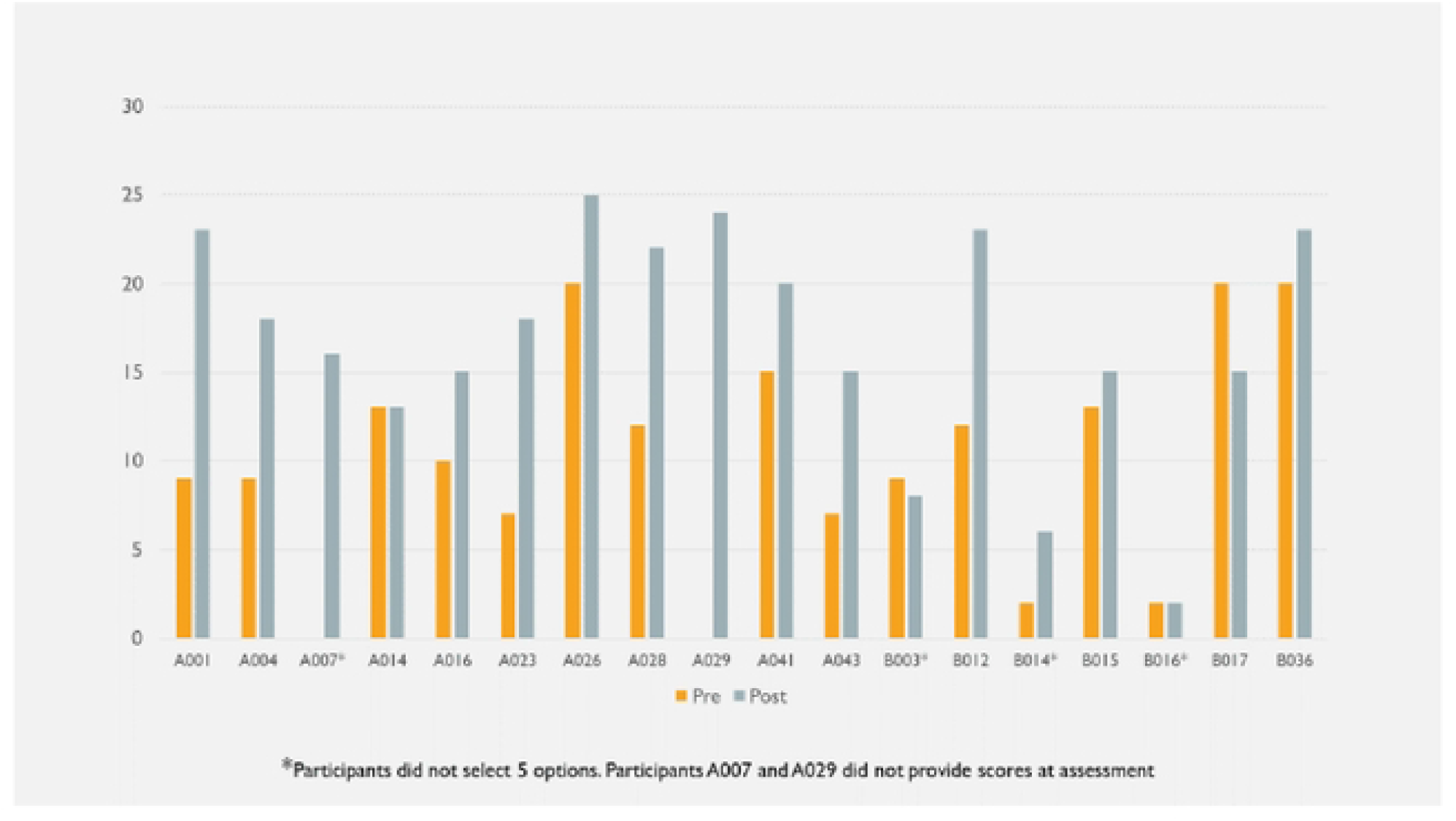
Assessment and follow up scores at Sites A and B.

### Interviews

Interviews were semi-structured, aiming to investigate further the responses provided in the surveys. For service users this was aimed at their own personal experience. For staff this aimed to cover all experiences of using the tool, including any use following the end of the study, with a focus on the practicalities.

### User interviews

Interviews were carried out with two service users in January 2021. Both were aged in their seventies and from organisation B/C. They stated that they had not had a previous wheelchair assessment before that in the study. These took place in March 2020, with equipment delivered in May 2020.

One was a woman who had been using a chair for three years following a stroke. On her survey she had responded positively toward the tool, stating it had taken 10 minutes to complete with assistance and that it was very helpful. The staff member completing their survey indicated it had added 15 minutes to the assessment. They had not rated usefulness but did comment that use had focused the assessment on the user’s needs.

The user’s key outcomes (satisfaction rating) were Safety (3), Pain and discomfort (3), Feeling included (2), Moving around (2), and Independence (2). Although she had not stated what she wanted for the last two on the tool, at the assessment and in the interview, she described feeling excluded, that people were looking down on her. Addressing the issues of wheelchair weight and an awkwardly positioned handbrake meant that she felt safer and far more positive about using her wheelchair:

> *“It’s a godsend, it really is, I’m not so frightened in it, as I was in the other one. “*

She felt the tool had been easy to use, relevant and didn’t feel anything needed changing.

> *“It was easy to understand. I could relate to what I used the wheelchair for…Especially the safety bit and how you feel in the chair*…”

The second was a man who had been using a wheelchair for two years post amputation. He was also positive about the tool itself, stating on the survey that it had been easy to understand and complete, although it had been filled in for him by his wife. The staff member’s survey indicated that use added 15 minutes to assessments but had assisted in setting goals and highlighted areas of importance to the client. The user’s key outcomes (satisfaction rating) were Activities (2), Independence (2), Moving around (1), Happiness (2) and Self-esteem (2). His wife was present during the interview and was invited to contribute by the user. He commented

> *“When we filled everything in for me [sic] wheelchair, until you’ve had it, you don’t really know what you do and don’t want”*

### Staff interviews

Interviews with staff included four clinicians who had used the tools in assessments themselves, including after the end of the study period: the manager/clinician from site A (*n*=20), the clinical lead for sites B/C (*n*=14) and a clinician from site B (*n*=11) and a clinician from site D (*n*=16). The PWB liaison officer from Site A who was responsible for administering the WATCh tools in advance prior to 35 assessments and the clinical operational lead from site C who had managed staff responsible for 14 assessments using the tool were also interviewed.

Overall they could see the benefits of using the tools to provide a structured approach and allowing the patient voice to be heard:

> *“Brilliant to take focus on and empower users – That is one of the real, real positives… gives them a voice*…*it’s about empowering our service users so they’re leading the assessment rather than being passive*…*” (Site B, clinician)*

The tools could assist discussion around PWBs although in some sites there was little experience of that:

> *“…feel it could be quite useful. The PWB process is to talk about options for them so they don’t have to go with our recommended provision… I can see there could be examples where it could be, for example if it picked up something that couldn’t be achieved with NHS provision, but I haven’t had that in practice to date*.*” (Site D, clinician)*

Potential issues in use were around the additional workload on staff and time taken to include use of the tool, particularly where staff were dealing with PWB implementation and in some cases COVID:

> *“… if the client didn’t turn up on time or was late then clinicians were finding that they were constrained anyway in clinic time, so having to sit and try and complete this tool before the clinic appointment then made their assessment rushed” (Site A, PWB liaison)*

While assessments with children and younger people went particularly well, older users and particularly those living in care homes had more difficulties:

> *“The people that seemed to show the biggest response was you know children and young people whose cognition was such that they can participate, they really valued I think being asked…. we had a thirteen-year-old in on Monday and he quite valued the process I would say” (Site B/C, clinical lead)*
>
> *“… residential homes, where there’s frequent staff working with that individual and they’re time limited with what time they can spend with that person… they weren’t very, engaging in it…” (Site D, clinician)*

Staff all considered that time was needed to embed the process into their own ways of working and in some cases the pandemic had affected this:

> *“…you just have to get comfortable so that it’s seen as a normal part of the assessment and not just an add-on…the WATCh documents have to be completed and then you find your own mojo as to how that’s incorporated into the assessment*.*” (Site B/C, clinical lead)*

Suggestions for ways to maximise efficiency in use of the tool were also sought. By the end of the study, sites reported providing the tools in advance where possible, rather than to the service user ‘blind’ in clinic. This gave service users and their carers time to consider which outcomes were of most importance to them and discuss them with others, and also reduced the time spent in clinic.

Issues raised by staff included concern that tools might raise unrealistic expectations about the equipment likely to be able to be provided, or about the benefits likely to be achieved. They noted that in a small number of cases, users did not engage with the tool as they ‘*just wanted a wheelchai*r’. Others considered some of the outcomes listed to be insensitive, for example, to a service user with a progressive or terminal illness. Some suggested that, where some patients struggled to select five options, three might be more appropriate. Some staff felt that the tools were more appropriate for new referrals than for users who were already familiar to the service.

## Discussion

There are several outcomes measures available for use by therapists and assistive technology providers in the UK, but at the time of developing the WATCh tool, none were specifically aimed at children and young adults requiring a wheelchair. The authors worked with children, young people and their carers to understand the wide range of outcomes from using their wheelchair that were important to them. The resulting tool aims to help wheelchair users and clinical staff identify key outcomes prospectively, and measure changes in outcome satisfaction after receiving new wheelchair equipment.

In 2014, Kenny and Gowran identified measures for wheelchair and seating provision which addressed an intervention’s contribution to the activity and participation of the individual and captured the influence of the entire service provision on the quality of life of the individual [12]. From their critical appraisal of 5 of the most promising measures, they concluded that no single outcome measure captured all necessary information and that trade-offs were inevitable.

Most measures in use assess patients using fixed predetermined areas, rather than focusing on the user’s own needs and preferences. For example, the Therapy Outcome Measures (TOM) [13], is used by rehabilitation professionals to assess patients against predefined areas using pre-coded levels of achievement. Adaptations of the TOM with specific levels of achievement have been developed for different clinical conditions, but none are focused on wheelchair use.

Other measures, aimed at users of wheelchairs also use predetermined areas with a focus on functionality. The Wheelchair Users Functional Assessment (WUFA) [14] evaluates ability to undertake a number of activities in an observed performance-based tool. The Functioning Everyday with a Wheelchair (FEW) [15] (also previously known as the Functional Evaluation in a Wheelchair instrument [16]), is a self-report scoring system on ability to carry out specific tasks. The Activities Score for Kids (ASK) self-completion measure also uses pre-determined items and focuses on functional ability [17]. The Functional Mobility Assessment (FMA) [18,19] obtains user ratings of agreement with statements related to predetermined functional areas.

In terms of assessing more qualitative aspects of equipment use, the Quebec User Evaluation of Satisfaction with Assistive Technology (QUEST) [20] evaluates levels of satisfaction with aspects of the service or the technology. While it is relevant to wheelchair users, it only captures satisfaction with what has already been provided. The Psychosocial Impact of Assistive Devices (PIADS) [21, 22], focuses on self-rated functional independence, well-being and quality of life of the patient. Initially developed for adults, it lists 26 predetermined areas for rating, while the children’s adaptation utilises a five-point Smiley Face Likert-type scale of agreement with 15 short statements to assess the constructs of Competence, Adaptability, and Self-esteem [23].

Other tools have aimed to determine outcomes defined by patients or in collaboration with patients but are not aimed at wheelchair users specifically. The Goal Attainment Setting (GAS) measure [24,25] is interview-based and can be used with clients with different problems and therapy approaches, to identify high priority goal areas, and agree specific and measurable indicators of progress. The Canadian Occupational Performance Measure (COPM) [26] is intended for use by occupational therapists with patients using semi-structured interviews to identify, rank in importance and rate performance and satisfaction with aspects of their life. Both have been used in paediatric research [27].

Acknowledging that there was “…no existing tool which can provide individualized goal-orientated measure of outcome after wheelchair provision”, led to the development of the Wheelchair Outcome Measure (WhOM) [28]. Clients nominate their key areas of participation inside and outside the home unprompted; rank their relative importance, and rate their level of satisfaction with each, at assessment and reassessment. It also includes a section for completion by the clinician. It has recently been adapted for use with young people as the WhOM-YP [29]. While the overall aim is very similar to that of the WATCh tools, the incorporation of clinical aspect may make it lengthier and more complex to deliver. Thus used as a whole it may not fit as an add on to the varying processes used by services. It may also be less suitable for sharing in advance to allow the client time to consider outcomes of importance to them.

Kenny and Gowran’s critical review identified several of the measures noted above and summarised their administrative burden, including the claimed time taken to complete. This ranged from up to 15 minutes for the QUEST and for the FEW, 30mins for WhOM and 1 hour for the GAS [12]. The PIADs is reported to take only 5-10 minutes although they cite a paper where use took 25 minutes. Thus the average time reported by staff and users to complete the WATCh and WATCh-Ad tools is in line with the tools that are quickest to complete, however it is acknowledged that the WhOM also incorporates some clinician-specific questions related to equipment provision.

The real-world study presented here gave an insight into how organisations adapted their use of the tools, alongside gaining familiarity with their use. Based on this, it is recommended that where possible, the WATCh tools and information on the PWB process should be provided alongside the invitation to the assessment appointment, whether paper-based or electronic.

The majority of responses from service users indicated that they had no problems or questions with the Tool. Assessors noted problems or questions in almost half of their survey responses, but the majority were resolved during the assessment. Particular issues arose with users who had problems with the written format, for example those without English as a first language, or who had issues with mental capacity, sight or literacy. Similarly, those with hearing difficulties had issues if the form was being read over the phone.

With regard to suggestions to reduce the number outcomes to be selected by users, it should be noted that even in this small-scale study, all options listed on the WATCh-Ad tool were selected by or on behalf of at least one service user, including the open ‘Other’ option. Similarly, in the pilot work undertaken as part of the development of the original WATCh for children’s tool, all items were selected by at least one child or their parent/guardian [7]. Whilst some of the reasons given in Part B could relate to a different area of Part A than was actually selected, this was not consistent enough to suggest any particular outcome listing was superfluous as it was covered by another. If anything, the reasons given around the most commonly selected ‘moving around’ outcome often related to one of the other areas and could be considered redundant. The tools are intended to assist service providers in identifying the outcomes of most importance to a user by prompting the user to think about wider aspects of their need for a wheelchair. Several of the users commented spontaneously that the list made them think of other aspects of their need for a wheelchair and enabled them to discuss these with the staff. As patient-centred rather than patient reported outcomes measures, the WATCh and WATCh-Ad tools obtain scores to be used for intra-individual comparisons of satisfaction before and after receipt of the equipment and are not intended to be used as a comparison between users, services or equipment. Where staff consider it appropriate or necessary in individual cases, selecting three outcomes might be sufficient to allow the assessment to focus on those most important to the user. The authors consider that wording the instructions to state that they can choose three to five options from the list might allow staff some leeway to discuss the user’s needs in more depth and focus down. It would still be possible to use the scoring tool to assess a percentage change in scores.

The type of assessment situation in which the tools should be implemented may need refining. Staff found them unnecessary in situations where straight-forward repairs or adjustments only were required. Early screening and triage of referrals and contact with users should be able to identify cases where the tool would not be necessary, and some individual service users or carers commented that staff already knew more about their needs and circumstances than tools could identify. However, as over three quarters of the users covered by the assessments stated that the tools were helpful or very helpful, this indicates that the tools helped them feel more involved in the decision-making even if the service’s choice of equipment was unchanged. We suggest that the tools should be offered to existing as well as new users, but that the accompanying information for users be worded to indicate that it is a standard approach being taken by the service, even for those users who were already well-known to them.

In this study the majority of service users providing feedback were positive about use of the WATCh tools in helping them consider their wider needs and discuss these with their wheelchair service provider. In addition, use of the tools had influenced the prescription and the PWB choice in up to 33% of cases at some sites.

The uptake of any new treatment or process is reliant on successful implementation. Proctor et al (2009) in work relating to mental health services in the United States referred to three kinds of outcomes in implementation research – client outcomes, service outcomes and implementation outcomes. Client outcomes were defined as satisfaction, function and symptomatology, with service outcomes based on the six Institute of Medicine (IoM) standards of Care published in 2001, namely efficiency, safety, effectiveness, equity, patient-centeredness and timeliness. Implementation outcomes were further developed into eight conceptually distinct outcomes of acceptability, adoption, appropriateness, feasibility, fidelity, implementation cost, penetration, and sustainability [30].

The WATCH and WATCh-Ad PCOMs tools are designed to uncover areas of importance to individual patients around satisfaction and function and to a certain extent symptomatology. This study looked at service outcomes plus the implementation outcomes of acceptability, appropriateness, and feasibility in order to make recommendations to maximising successful adoption, fidelity, uptake and sustainability. Further work is required to fully assess the relationship between costs and benefits of implementing a PCOMs / PWB process on wheelchair provision.

The study also allowed assessment of the aspects of validity relevant to PCOMs [31] which by definition are used for intra-person evaluation over time and not intended for use in a comparative test: content validity, face validity and clinical validity.

Content validity relates to the ability of a tool to measure what it is supposed to measure. The initial WATCh tool was developed through previous academic work with children/young people and based on work on quality of life in people with mobility impairments by the authors and was deemed relevant to be adapted for use with adults. Although some participants in this study felt the number of items was too long, all outcomes listed were selected as important by at least one participant.

Face validity refers to the acceptability to the test taker. The present study added information on adult service users and their carers to the information gained from the work developing the WATCh tool for children [7, WATCh], and found that use of the tool was considered helpful or very helpful to a majority of participants. However further work is needed to increase usability among service users with additional needs. Ease of use could be increased further by enhancing the ability for the tools to be shared between service user and the service by electronic means.

Clinical validity is whether a measure has overall usefulness in a clinical situation. Although in this study there was a slight majority of assessments where staff did not feel that the tool had been helpful or had altered the prescription, an impact was noted in 24%.

The authors acknowledge potential limitations of the work. This was a relatively small study which aimed to review the use of the tools in varied situations, within a short timescale. From a practical perspective, there was not time to formally pilot the survey tools, however the study team, including clinicians, commissioners and a service user reviewed the protocol and documents. In hindsight it would have been preferable to include more options around the level of usefulness to staff in place of a yes/no option.

As described above, the impact of the COVID-19 pandemic on the NHS in terms of capacity and ability to perform research affected the timing and thus the numbers of assessments able to be reported upon, and in particular the ability to obtain more in-depth views from the service users. Although many of the assessments at sites A to C took place before the announcement to stop providing all but emergency provision in the NHS, there was significant media interest in the pandemic and those attending may have been the most severe or urgent cases. After the restart there were still significant concerns about the risk of infection, and this may have compounded differences between Site D and the other sites based on type of service or stage of experience of use of PWB or the WATCh tool. There was reluctance by some to attend clinic or have visitors to the home. Indeed, the number of patients registered, and the number of new referrals for both adults and children recorded in the latest available data [2] has decreased from the numbers reported in October-December 2019, the last quarterly data obtained before collection was paused [32]. At that point there were over 700, 000 registered users including over 75,000 children, and an average of over 300 new referrals per quarter per CCG.

Measures taken to reduce transmission of the virus also led to implementation of different ways of working, for example, making more use of telephone screening and/or digital technology for screening/triage and digital technology for remote assessment and use of home visits. Communication via smart phone or email was introduced to speed up the time for appointment notifications to be received by service users. However, there is evidence that some of these new ways of working may be in use for longer than just the immediate period of the pandemic, hence information gained on benefits and issues relating to use of the WATCh tools alongside these in this study will remain relevant going forwards.

It was hoped that the use of such tools would increase the ability for the voices of the service users to be heard. Due to the circumstances of many service users, responses were obtained through the carer and in some cases with the help of the assessor. The original WATCh tool was developed for children to complete themselves. In this study, while the results from the staff interviews noted that the tools were generally well received by children and young adults, all the survey respondents completing the WATCh tool for children were parents or carers. In many cases there will have been practical, physical or capability reasons why a child could not respond for themselves, but it will be important that assessors ensure that children and young people, and indeed adults where a carer has provided the responses, agree with the outcomes selected on their behalf as far as possible.

The addition of any new practice into an existing way of working would be expected to increase the time involved, at least until a service assesses the most efficient ways of incorporating it into standard practice. This can be achieved through staff training and mentoring, and by encouraging staff to develop their own way of working with it within the protocol. Practical suggestions noted above include providing the tool in advance with appropriate instruction and developing ways of using the tools alongside new digital technologies which are already increasing in part due to the pandemic. Further work is needed to broaden accessibility and service utility of the tools, for example translations, digital availability and automatic linking of tool scorings into the service user’s record.

## Conclusions

The majority of participating wheelchair service users stated that the WATCh and WATCh-Ad patient centred outcomes tools were helpful, assisting them in identifying their required outcomes and discussing these with staff at their assessments. Staff noted that use of the tools impacted on prescriptions or assisted in the choice of PWB in just under a quarter of their assessments, a finding which if rolled out nationally would extrapolate to a large number of service users. Concerns were expressed by some about the time required to incorporate the tools into standard assessment practices. The COVID-19 pandemic severely reduced the time and local resource available within the study period for use of the tools to become fully embedded in practice, as required for implementation of any new process particularly at the sites with no prior experience of them. However, it was sufficient to allow for a number of new approaches to be implemented to increase the usefulness and efficiency of the Tools, such as provision of the tools in advance of appointments to allow users time to consider their needs and reduce the time in clinic. Further enhancements should include availability in digital format and ultimately the ability to auto-populate existing clinical systems.

## Data Availability

Data cannot be shared publicly as the authors consider that the small number of specialised sites in discrete geographic areas, combined with the timing of attendance and information on underlying problems could uncover anonymity. Data will be available on reasonable request from the corresponding author.

## Acknowledgments

The authors wish to thank the study team and the participating staff and service users at the sites involved, along with the support of Hull CCG, East Riding of Yorkshire CCG, Blatchford Limited, NRS Healthcare, and Dr Catherine Lawrence, Bangor University, for reviewing the manuscript.

## Supporting Information

**S1 Appendix: WATCh-Ad Assessment and follow up tools as used**

**S2 Appendix: WATCh Assessment and follow up tools as used**

**S3 Appendix: Staff and User Surveys**

## Abbreviations

CCG: Clinical Commissioning Group
COVID-19: COrona VIrus Disease 2019
EoI: Expression of Interest
GDPR: General Data Protection Regulations
IoM: Institute of Medicine
LREC: Local Research Ethics Committee
NHS: National Health Service
NIHR: National Institute for Health Research
n/s: Not stated
NWAG: National Wheelchair Advisory Group
OT: Occupational Therapist
PCOM: Patient Centred Outcomes Measure
PT: Physiotherapist
PWB: Personal Wheelchair Budget
R&D: Research and development
RE: Rehabilitation Engineer
SU: Service user (term used in responses)
WATCh: Wheelchair outcomes Assessment Tool for CHildren
WATCh-Ad: WATCh tool for ADults

